# Toward Digital Twins in the Intensive Care Unit: A Medication Management Case Study

**DOI:** 10.1101/2024.12.20.24319170

**Authors:** Behnaz Eslami, Majid Afshar, Samie Tootooni, Timothy A. Miller, Matthew M. Churpek, Yanjun Gao, Dmitriy Dligach

## Abstract

**Objective:** To evaluate the efficacy of digital twins developed using a large language model (LLaMA-3), fine-tuned with Low-Rank Adapters (LoRA) on ICU physician notes, and to determine whether specialty-specific training enhances treatment recommendation accuracy compared to other ICU specialties or zero-shot baselines.

**Materials and Methods:** Digital twins were created using LLaMA-3 fine-tuned on discharge summaries from the MIMIC-III dataset, where medications were masked to construct training and testing datasets. The medical ICU dataset (1,000 notes) was used for evaluation, and performance was assessed using BERTScore and ROUGE-L. A zero-shot baseline model, relying solely on contextual instructions without training, was also evaluated. While our approach moves toward digital twin capabilities, it does not incorporate real-time, patient-specific EHR data and can be viewed as an ICU specialty-level language model adaptation.

**Results:** Models fine-tuned on medical ICU notes achieved the highest BERTScore (0.842), outperforming models trained on other specialties or mixed datasets. Zero-shot models showed the lowest performance, highlighting the importance of training.

**Discussion:** The findings demonstrate that specialty-specific training significantly improves treatment recommendation accuracy in digital twins compared to generalized or zero-shot approaches. Tailoring models to specific ICU domains strengthens their clinical decision-support capabilities.

**Conclusion:** Context-specific fine-tuning of large language models is crucial for developing effective digital twins, offering foundational insights for personalized clinical decision support.

## INTRODUCTION

A digital twin is a virtual model, continuously synchronized with real-time data, that replicates a physical object, system, or process. By integrating diverse data sources and leveraging predictive analytics, it enables simulation, monitoring, and analysis of its physical counterpart, fostering improved decision-making and optimization.^1^ A digital twin in the medical context is a dynamic, data-driven representation of real-time clinical scenarios that evolves in response to patient data and advancing medical knowledge.^2^

Functioning as a bridge between healthcare providers and artificial intelligence systems, such as large language models (LLMs), a digital twin continuously adapts to the types of patients seen by a provider as electronic health records (EHR) expand with new data and disease insights. By integrating its internal medical knowledge with EHR data, the digital twin can assist in diagnosis and treatment reasoning, helping to alleviate the cognitive burden and information overload often encountered in the care of complex critical care patients.^3,4^ Digital twins can be tailored for varying levels of granularity, ranging from a single provider to an entire medical specialty or healthcare organization, offering scalable and personalized support across diverse clinical settings.

In critical care medicine, a digital twin can have a particularly high impact given the dynamic and time-sensitive nature of intensive care units (ICU), drawing on massive amounts of data that are often beyond the processing abilities of an individual physician. The typical ICU is not a monolith, but rather a consortium of multi-disciplinary specialists with different training paths and experience. For example, a patient with a major cerebrovascular accident would be treated in the Neuro ICU, a severe trauma in the surgical ICU, and septic shock in the Medical ICU. Our choice of the level of granularity at which we perform our modeling is driven by the availability of training data; we resort to the Medical Information Mart for Intensive Care III (MIMIC-III) corpus^5^ of over two million critical care notes that can be further stratified into several ICU specialties (**Figure 1**), facilitating the creation of ICU specialty-specific digital twins.

**Figure 1.**
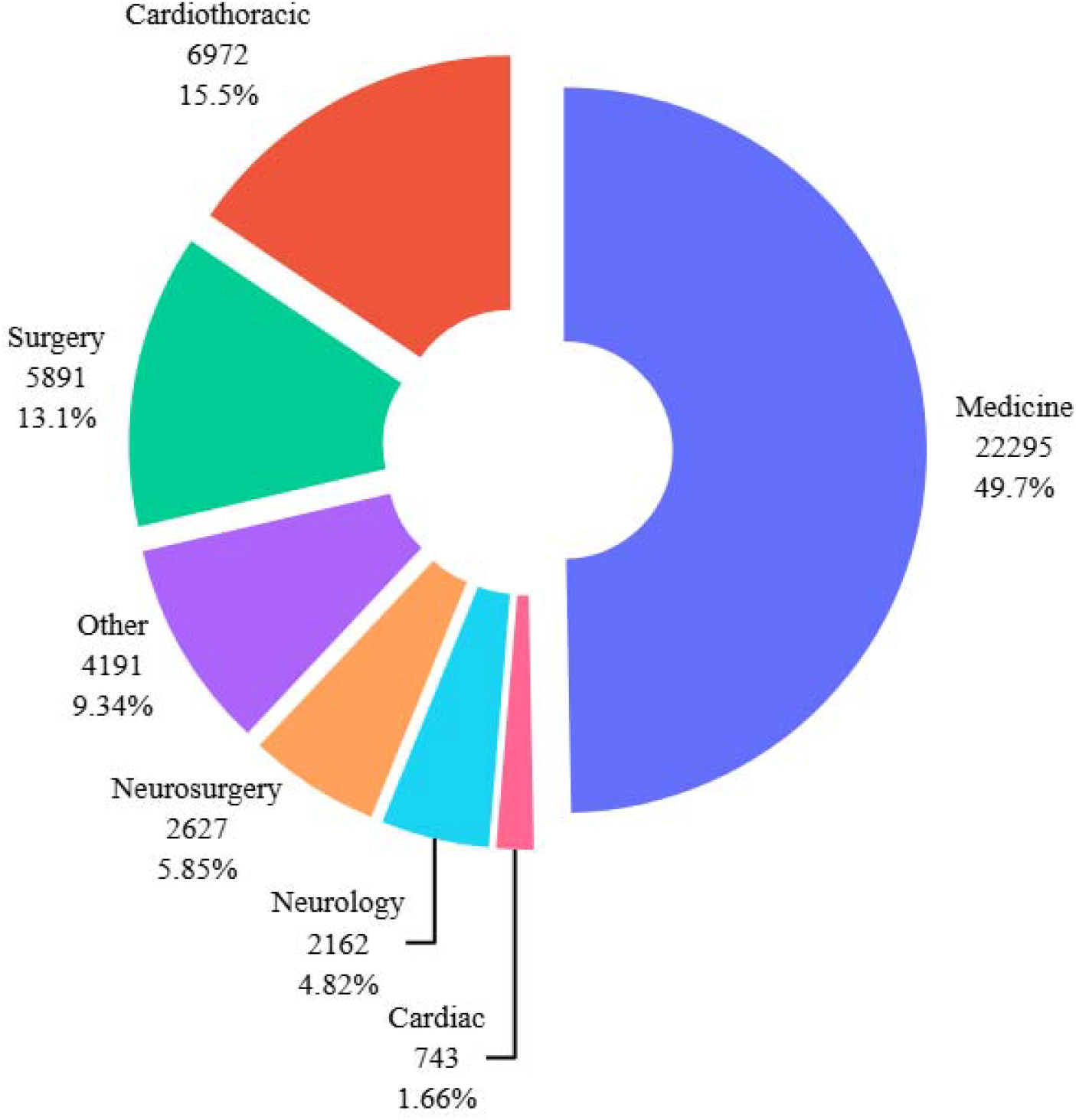
Frequency Distribution of Service Types in MIMIC-III Note Events

In this work, we use the term “digital twin” in a domain-specific, functional sense: as a surrogate model that reflects the treatment behaviors and preferences of ICU specialty providers based on historical patient data for a given ICU disease area. While our implementation does not involve real-time synchronization with EHRs - a characteristic often emphasized in industrial digital twin applications^6,7^-it retains the core properties of a digital twin by enabling simulation, reasoning, and adaptation based on specialty-specific clinical data. Specifically, we frame our work as an offline simulation task that mimics the treatment decision processes within different ICU specialties. The “twin” in this context is not an individual patient, but a data-driven model of the ICU specialty itself - capturing its evolving clinical patterns. As such, our approach aligns with a growing class of conceptual or knowledge-based digital twins, which aim to replicate decision-making behaviors through machine learning rather than physical or real-time system modeling^8^ The potential application of an ICU specialist represents the differing diseases seen between ICUs. For example, a patient in shock could be due to hemorrhagic shock from intrabdominal trauma, septic shock from pneumonia, or cardiogenic shock from acute coronary syndrome. The management and treatment of each type of shock patient requires a specialist intensivist with training and expertise in that disease area.

We begin by introducing a new and challenging modeling task: ICU medication prediction. The total number of *unique* medications mentioned in MIMIC Medical ICU discharge summaries exceeds 14,000, making the task challenging for both humans and machines. The task was designed as a sequence generation task where the input to an LLM are sections from the discharge summaries with *all* medication mentions masked using a special token. The generated output from the LLM was a prediction of the masked medications. A synthetic example that illustrates this task is shown in **Figure 2** (our data use agreement with PhysioNet does not allow us to include actual MIMIC notes).

**Figure 2.**
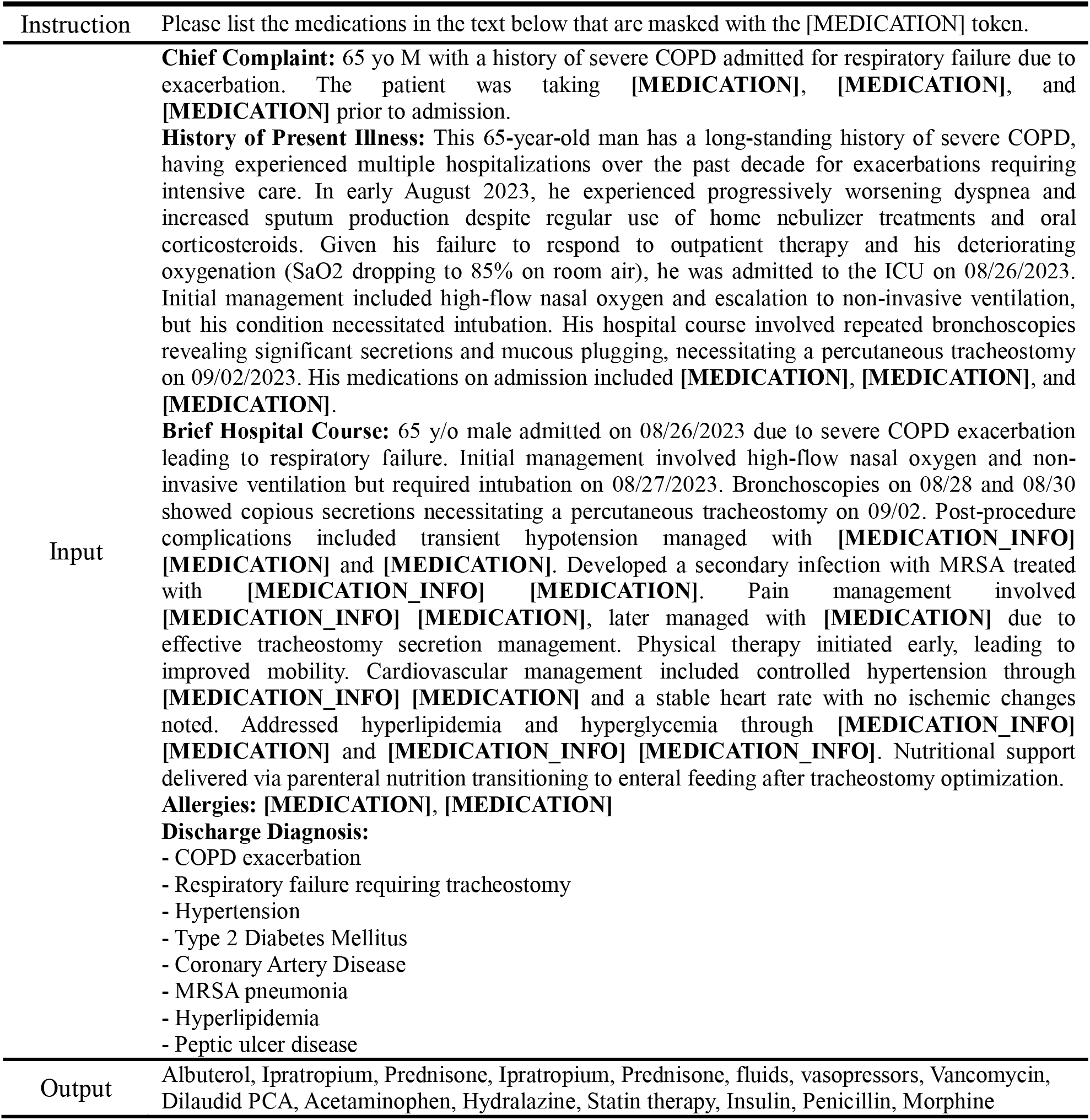
Sample synthetic note with masked medication mentions like the notes used for supervised fine-tuning. We train LoRA adapters using thousands of similar notes to predict the identity of the masked medication mentions.

Best practices for common ICU medications like vasopressors or sedation often vary between ICU specialties. We proceed by training digital twin classifiers on notes from different ICU specialties and assessed their performance on specialty-specific notes, comparing zero-shot learners with LLMs adapted to distinct ICU specialties to measure shifts in treatment preferences. Open-source LLMs were used in zero-shot experiments to represent a physician without any knowledge of preferences that may exist within an ICU specialty. The digital twin was trained by adapting an LLM, LLaMA3^9^, to reflect treatment preferences specific to an ICU specialty using Low-Rank Adapters (LoRA)^10^. For example, we used notes written by physicians working in a cardiothoracic ICU to train a cardiothoracic digital twin.

Unlike some digital twin implementations reported in the literature, our current approach does not involve real-time EHR data integration due to the static nature of the publicly available dataset used for model training. As such, our work should be viewed as an ICU specialty-level language model adaptation rather than a fully realized digital twin. However, enabling real-time integration remains a promising direction for future research. Our use of Low-Rank Adaptation offers a computationally efficient pathway toward such integration compared to full-parameter model training.

We evaluated our digital twins by measuring their performance on the notes from the most common ICU type, Medicine. Our hypothesis was that there are measurable differences across the treatment courses that are used across different ICU specialties and an LLM adapter can be trained to capture those preferences, effectively creating a digital twin. The datasets are available to the community to facilitate future work on this task. The full implementation, including data preprocessing scripts and model training configurations, will be available at [GitHub link] upon acceptance.

## RELATED WORK

Digital twins in healthcare traditionally represent virtual counterparts of patients, aiding in simulating outcomes, forecasting treatment responses, and advancing precision medicine.^11^ Vallée et al.^3^ demonstrates the potential of digital twins for real-time monitoring and predictive analytics in chronic disease management, highlighting their role in providing proactive and individualized care. Our approach extends this concept to provider-specific digital twins that adapt to ICU specialties, incorporating both clinical expertise and local practice behaviors captured in the EHR. LoRA adapters proved instrumental in this task, offering efficient fine-tuning with fewer parameters and enabling models to reflect provider-specific practices. This adaptability is critical, as ICU specialties vary significantly in their treatment protocols and clinical priorities. Additionally, the flexibility of LoRA allows for dynamic updates, ensuring that digital twins remain current with evolving EHR data and guidelines.

LLMs like GPT-4^12^ and LLaMA^13^ have shown great promise in medical document processing^14^, becoming increasingly important in medicine and medical informatics by enhancing diagnostic precision and treatment decisions through data-driven insights^15^. Our experiments indicated that LLMs can be trained to reflect the differences in treatment methods across different ICU specialties, similar to work by others. Liu et al.^16^ proposed a framework where LLMs, fine-tuned with a prompt template, train a smaller student model via knowledge distillation to recommend medications by adjusting output probabilities; unlike their approach, our method uses LoRA to adapt LLMs to ICU specialty-specific data, emphasizing domain-specific treatment behaviors rather than model compression. Moreover, Dou et al.^17^ introduced ShennongGPT, an advanced LLM designed for medication guidance and adverse drug reaction prediction, using a two-phase training approach with drug databases and real-world patient data; in contrast, our work captures practice-based treatment patterns rather than relying on structured pharmacological resources. Additionally, Ahmed et al.^18^ leveraged the latest advancements in LLMs for domain-specific factual knowledge, presenting MED-Prompt — a novel prompt engineering framework designed to achieve accurate medicine predictions; in contrast, our approach through fine-tuning LLMs using LoRA, emphasizing adaptation to real-world specialty practice patterns rather than prompt optimization alone.

The growing evidence supports using LLMs as a foundation for creating tailored digital twins in critical care settings.

## MATERIAL AND METHODS

### Data Corpus and Study Setting

MIMIC-III database is an open-access resource that provides de-identified health data from over 40,000 patients treated in intensive care units, including free-text admission notes that detail patient encounters.^19^ For this study, we obtained the counts of discharge summaries in MIMIC-III across different ICU specialties (**Figure 1**) and selected the three most frequent ones: (1) Medical ICU; (2) Cardiothoracic ICU; and (3) Surgical ICU. This study was reviewed and determined to be exempt from oversight by the Loyola University Chicago Institutional Review Board (IRB #3834, Application #10225). Using a simple rule-based approach, we identified the five sections in the discharge summaries that are most relevant to clinical care during the hospital course: (1) Chief Complaint (CC); (2) Brief Hospital Course (BHC); (3) History of Present Illness (HPI); (4) Allergies; and (5) Discharge Diagnosis (DD). Sections were identified using regular expression matching based on commonly occurring section headers in discharge summaries, such as “Chief Complaint:”, “Brief Hospital Course:”, “History of Present Illness:”, “Allergies:”, and “Discharge Diagnosis:”. Text corresponding to each section was extracted until the next known section header was encountered. These sections are ubiquitous to discharge summaries at many health systems and contain the most detailed content of the hospital and ICU course, including the medications prescribed.

SparkNLP^20^ was applied to automatically identify all medication mentions in the discharge summaries. A critical care physician and clinical informaticist (MA) manually reviewed 20 randomly selected notes and confirmed high medication detection accuracy, with SparkNLP achieving an average F1 score of 0.944 across medication mentions and their associated attributes, including dosage, strength, route, frequency, and duration. Further details of this evaluation are provided in Table S1. ^20^For each note, the tagged medication mentions were masked with the special token [MEDICATION] (see **Figure 2** for a sample note). In addition to medication mentions, SparkNLP tagged other medication-related entities: DOSAGE, STRENGTH, ROUTE, FORM, FREQUENCY, and DURATION. To avoid revealing the identity of the masked medication, the additional entities were masked with a special token [MEDICATION_INFO]. The total number of unique medications for each ICU specialty, along with the top 10 medications and their frequencies, are shown in **Table 1**. The masked medication mentions were retained and used as the reference labels for the prediction targets. This preprocessing resulted in a dataset of 18,830 Medical ICU notes. To create our training and evaluation data, each note was paired (with all medication mentions masked) with a comma-separated list of medication mentions from that note that served as the prediction targets. The dataset was split into training (16,330), development (1500), and test (1000) sets. The details of the training, validation, and test corpora are provided in Table S2 and Table S3. Additionally, the demographic and clinical characteristics of the patient cohort are summarized in Table S4. An additional four training sets were curated, each containing 4,118 notes, for the most common ICU specialties: Surgery, Medicine, and Cardiothoracic, and a random sample from all 3 ICU specialties. The dataset was split at the note level. Although a small number of patients (<0.1%) appeared in more than one split, we deemed this overlap negligible for the purposes of this study.

**Table 1:**
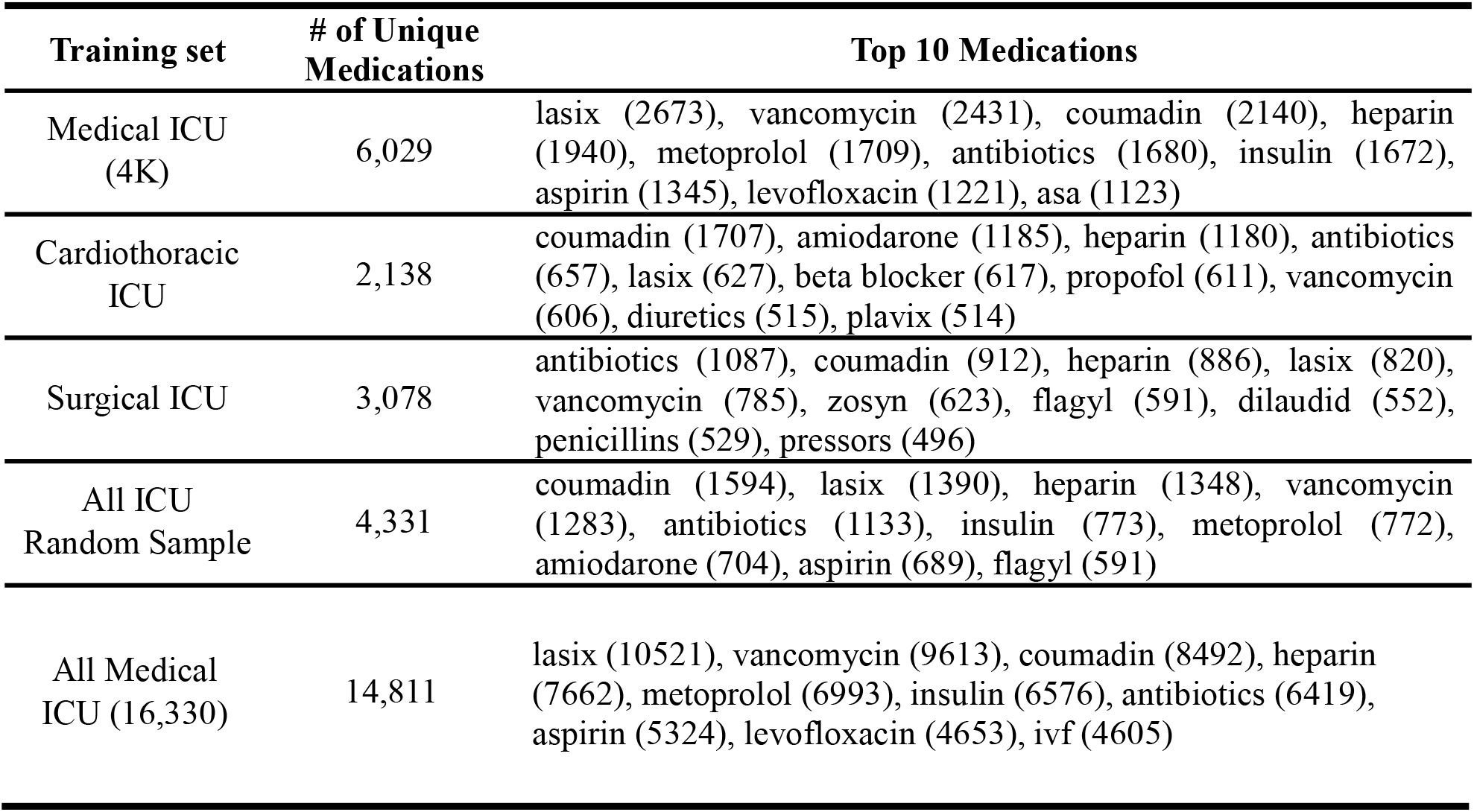
Unique Medication Counts and Frequencies of Top 10 Medications. The high number of unique medications mentions in discharge summaries makes our medication prediction task very challenging.

### Experiments

#### Supervised fine-tuning (SFT)

Llama3 LLM (llama3-8B-Instruct)^21^ was fined tuned using a LoRA adapter to accept notes from a single type of ICU and predict all medication mentions that were masked. This method leveraged low-rank factorization, reducing the number of trainable parameters and mitigating overfitting risks. We performed an extensive hyperparameter search to optimize the training and LoRA adapter parameters to ensure stable and efficient model training. After completing the model selection step using our validation set, a final evaluation was conducted using the test set, which also consisted of Medical ICU notes, and reported the results in **Table 2**. The average training runtime across the five corpora was approximately 5 hours, utilizing 8 NVIDIA RTX A6000 GPUs, each with 48 GB of RAM. The longest runtime was observed for the “All Medical ICU [16,330 notes]” dataset, which took approximately 13 hours.

**Table 2.**
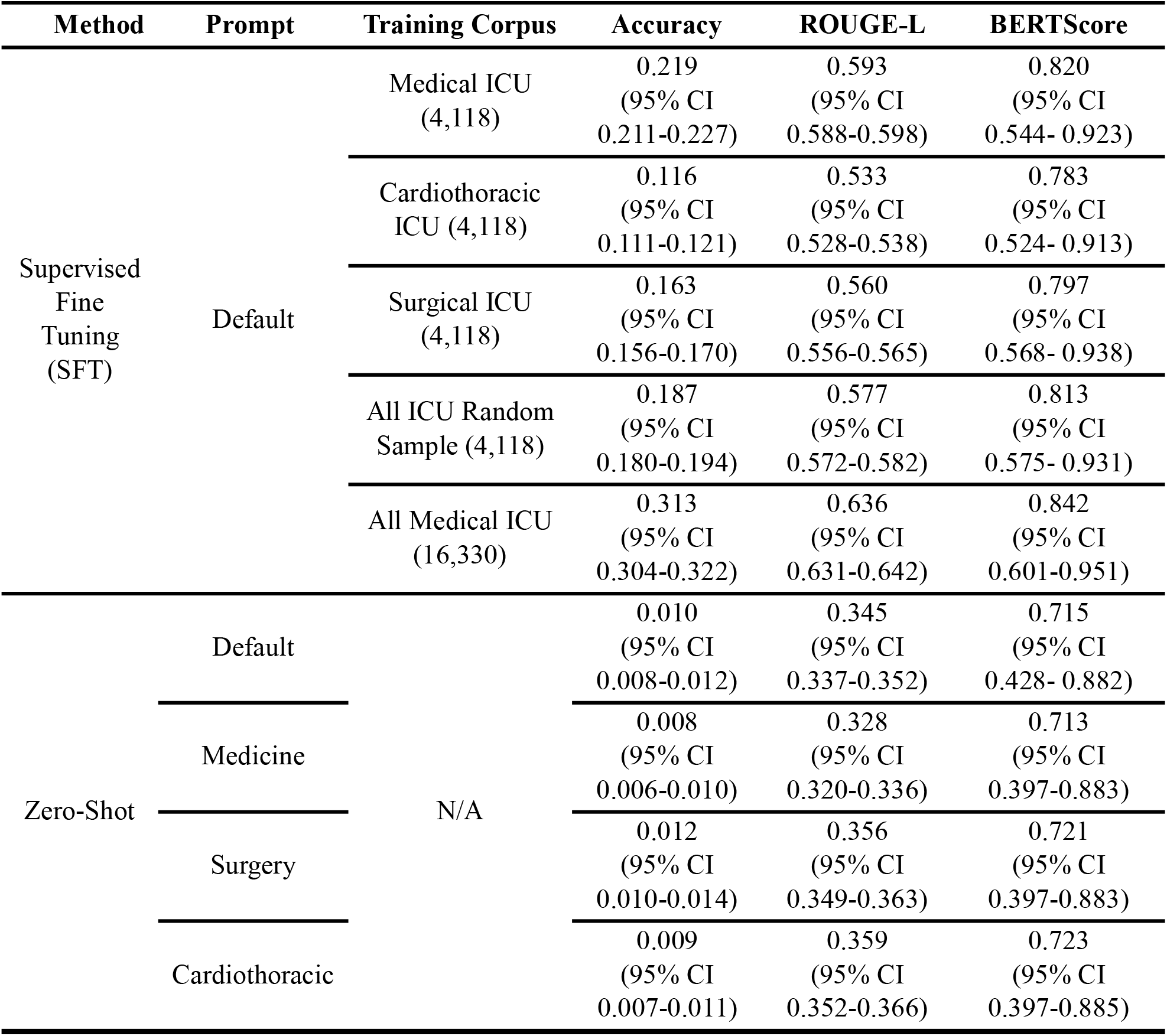
Medication prediction performance on Medicine notes for supervised fine-tuning (SFT) and Zero-shot learning experiments. For SFT, LoRA adapters were trained to reflect medication preferences within a single ICU specialty. For zero-shot learning, prompts were designed for an ICU specialty-agnostic prompt (“Default” prompt above) and ICU-specific prompts (e.g. a prompt that begin with “You are a Cardiothoracic ICU physician …”). The accuracy was measured as the proportion of exact matches between predicted and ground truth medications, and the 95% confidence intervals were calculated using bootstrap resampling with 10,000 iterations.

#### Zero-Shot Learning

To understand better how well the supervised models captured the preferences of ICU physicians from a given specialty of ICU, the fine-tuned models were compared to a model that did not have access to ICU-specific training data using zero-shot learning. Using the same LLM as in SFT, the prompt for zero-shot was refined using the development set. The final prompt is shown in **Figure 3**. Sequential numbering was applied to the masked medication entities within each clinical note to assist the model in predicting the correct number of medications. Additionally, we tested whether zero-shot learning can induce ICU-specific behavior by prepending to the prompt the sentence “You are a [ICU_specialty] ICU physician.”, where [ICU_specialty] is Medicine, Cardiothoracic, or Surgery. Following the generation of results, we conducted a post-processing step to remove any extraneous elements, thereby refining the final output for more accurate evaluation. The evaluated hyperparameter values for SFT, along with the optimal configurations highlighted in bold, are provided in Table S5.

**Figure 3.**
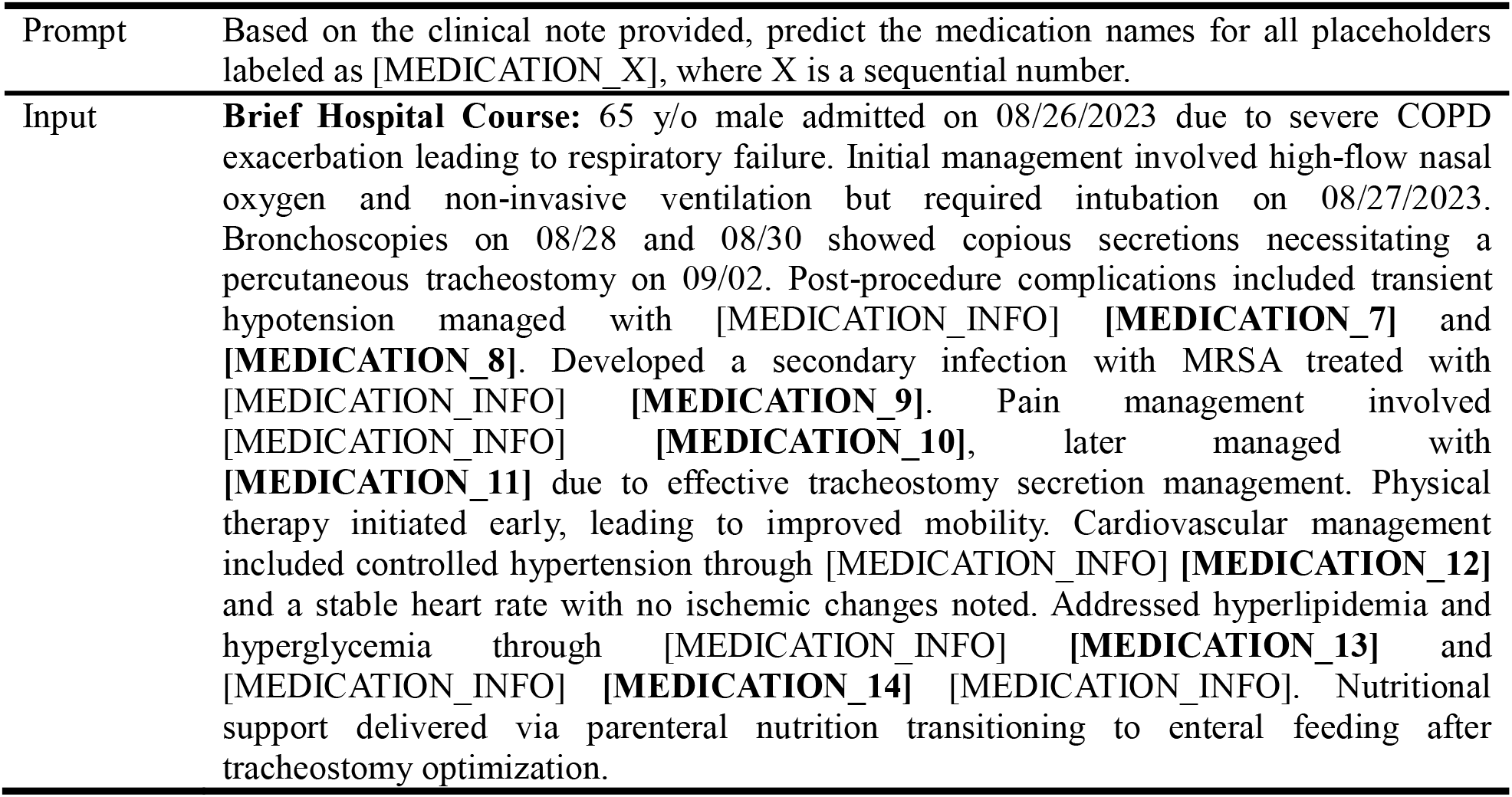
A sample synthetic note, similar to the notes used for zero-shot learning. An LLM is given a prompt that asks to identify the masked medication mentions.

#### Evaluation metrics

Evaluating the performance of the models presented several challenges. First, some medication names were synonyms and were used interchangeably (e.g., “ibuprofen” and “advil”). Next, medication classes were frequently substituted for specific medication names (e.g., “NSAID” instead of “aspirin”). Finally, there was no guarantee that the length of the generated sequence of medication mentions would match the length of the reference sequence. To address these complexities, we employed multiple evaluation metrics. First, the exact match accuracy was calculated by assuming a one-to-one alignment between the generated and reference sequences. The assumption of one-to-one match was violated in at least 20% of the cases and thus the exact match accuracy likely underestimated the model’s performance. The accuracy was calculated as the proportion of exact matches between the predicted medication sequences and the ground truth sequences. To get a better estimate of the model performance, we opted for two metrics that are routinely used for evaluating generative models: BERTScore and ROUGE-L. BERTScore is a soft match metric, with SapBERT^22^ as the base encoder. We chose SapBERT because it was trained on the Unified Medical Language System (UMLS)^23^ and had a strong correlation with human judgments.^24^ In our experiments, we found that BERTScore exhibited a high variance between longer and shorter sequences.^25^ This limitation highlighted the need for complementary metrics; therefore, we also reported ROUGE-L, which provided a more balanced assessment by focusing on the longest common subsequence, thereby mitigating the problem of aligning sequences. Our primary evaluation metric was the exact match accuracy score, and we used it for model selection. We compute 95% confidence intervals using bootstrap resampling with 10,000 iterations.

## RESULTS

Training data consisted of five datasets of ICU notes, as detailed in **Table 2**. For evaluation, the development and test sets included only Medical ICU notes, with 1,500 notes allocated to the development set and 1,000 notes to the test set. The performance of various models was assessed using accuracy, with the results on the test set of Medical ICU notes presented in **Table 2**. The flow diagram and final set of experiment architectures are shown in **Figure 4**. Models trained on the Medical ICU specialty demonstrated the best performance with a Bidirectional Encoder Representations from Transformers Score (BERTScore)^26^ of 0.842, achieving markedly better results than those trained on the notes from other ICU specialties. Even the training scenario that used all ICU specialties had a lower BERTScore at 0.783. The Cardiothoracic ICU digital twin had the worst performance, revealing a distinct gap between physician preferences for medication use in Cardiothoracic versus medical ICU specialty.

**Figure 4.**
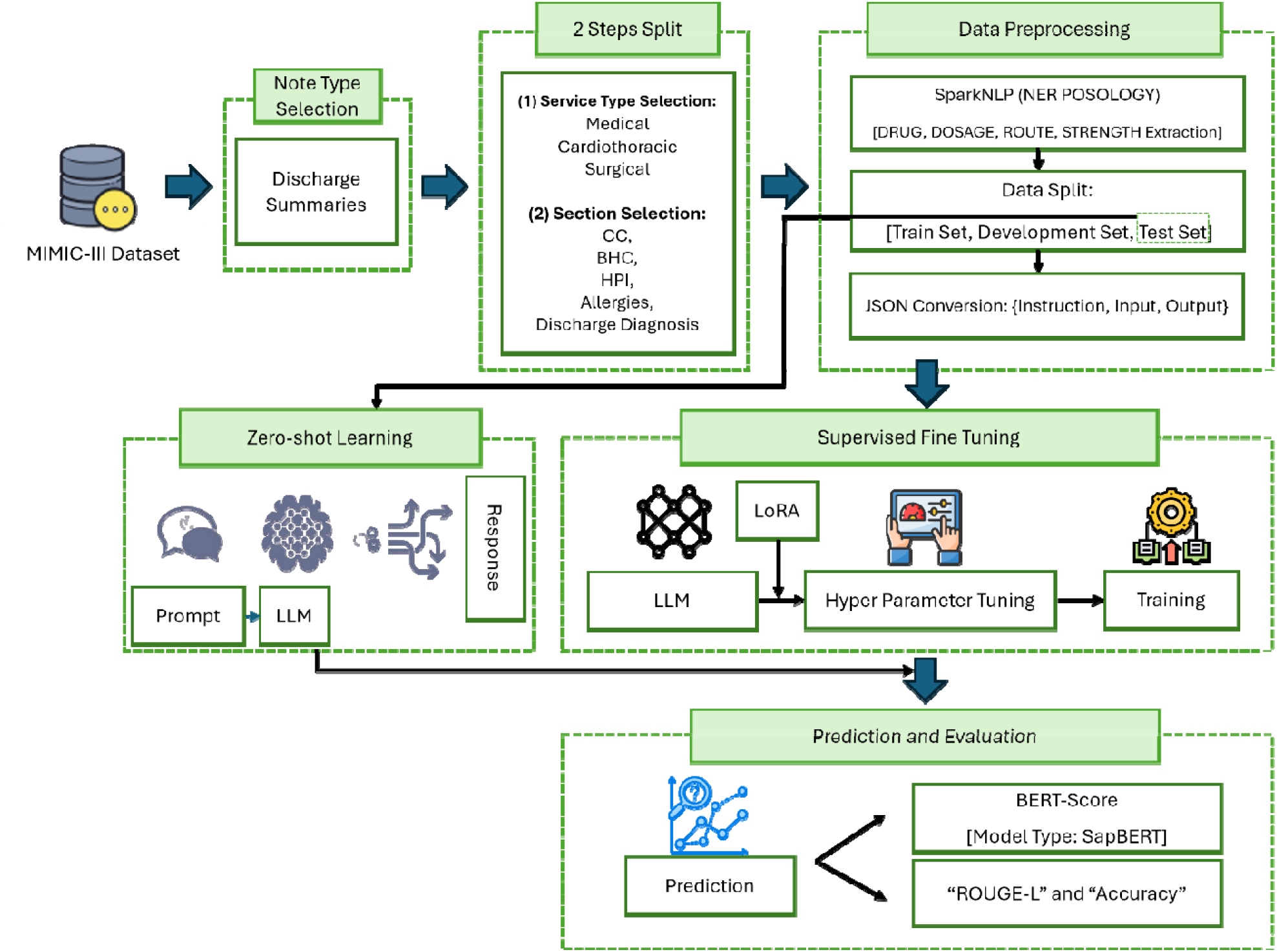
LLMs Medication Prediction Flow Diagram

The zero-shot models performed substantially worse, particularly in terms of accuracy, which was much lower than other models. We attributed this performance gap to challenges in designing prompts that effectively extracted medication names, as demonstrated by instances where the model returned medication categories (e.g., “analgesics” instead of “dilaudid,” or “antibiotic” instead of “ceftriaxone”). This was supported by a BERTScore of 0.713 for the zero-shot model that was closer to the performance of the supervised models but lower than the other models. Customizing the zero-shot prompt to specific ICU specialties, as shown in the bottom three rows of **Table 2**, did not improve the performance compared to the default (ICU-agnostic prompt), highlighting the need of supervised learning for creating digital twins.

While the medical ICU digital twin achieved relatively high performance with the BERTScore, it gave partial credit to approximate matches such as “ibuprofen” and “NSAID,” and in some scenarios such credit may not be warranted. While BERTScore effectively measures semantic similarity between two sequences, its inherent variability with longer sequences — likely influenced by the high variance in sequence lengths across notes — underscores the importance of utilizing n-gram overlap metrics like the Recall-Oriented Understudy for Gisting Evaluation - Longest Common Subsequence (ROUGE-L).^27^ ROUGE-L provides a more stable assessment of token-level overlap and alignment, ensuring a comprehensive evaluation of model performance despite differences in note length. Our results remained consistent when examining ROUGE-L scores and accuracy, with the medical ICU digital twin model trained on all medicine notes performing the best **(Table 2**).

To assess variability around the point estimate scores, 95% confidence intervals were calculated using 1000-iteration bootstrapping. The BERTScore showed high variance as demonstrated by the wide confidence intervals across all scenarios. To further examine the variance, the individual confidence intervals across different medication sequence lengths generated by the LLM are shown in **Figure 5**. Shorter medication sequences were associated with greater BERTScore variability, reflected in wider confidence intervals, indicating higher uncertainty in the score estimation. Conversely, longer sequences resulted in narrower confidence intervals, suggesting more robust and consistent BERTScore measurements.

**Figure 5.**
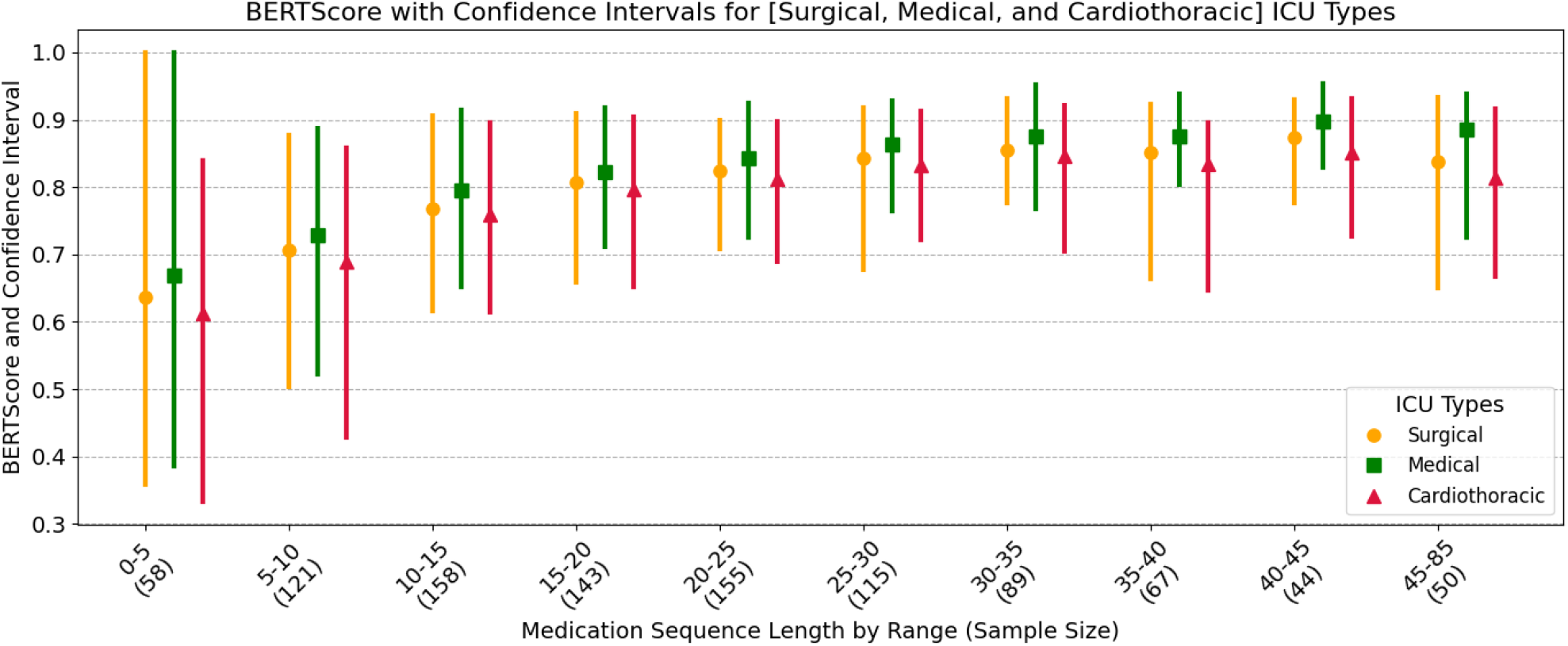
BERTScore with confidence intervals for different ICU specialties across medication sequences of different lengths binned into 10 intervals. The number below each interval is the sample size for that range. Overall, shorter sequences are associated with wider confidence intervals. Longer sequences, on the other hand, have tighter confidence intervals.

The variability in confidence intervals for sequence length in generated medications is an inherent characteristic of BERTScore. This metric relies on pairwise semantic comparisons, and as the number of available medication entities increases, the likelihood of identifying closely matching pairs rises. Consequently, longer sequences enable more comparisons, contributing to statistical variability in the BERTScore. In at least 20% of the test set notes, the models generated medication sequences did not match the reference medication sequence lengths **(Table 3)**. The number of mismatched sequences were as high as 55% for the model trained on cardiothoracic ICU notes.

**Table 3.** Percentage of test notes for which the number of generated medication sequences does not match the number of reference medications.

| ICU Specialty | Notes with mismatched length, % |
| --- | --- |
| Medical ICU (4,118) | 22.2 |
| Cardiothoracic ICU (4,118) | 55.1 |
| Surgical ICU (4,118) | 21.4 |
| All ICU Random Sample (4,118) | 20.4 |
| All Medical ICU (16,330) | 7.9 |

### Error Analysis

To better understand the types of errors made by our models, we conducted a manual error analysis. A physician expert (MA) reviewed 100 randomly selected validation notes — 20 from each of the five specialty fine-tuned models. The total number of predictions examined was 100. Errors were categorized into six types based on the nature of the incorrect prediction, and their relative frequencies are summarized in Table 4. Notably, the majority of errors (categories 3 and 4) involved medications that were either contextually appropriate but incorrect, or both inappropriate and incorrect, highlighting the challenge of medication prediction in complex clinical narratives.

**Table 4.** Distribution of prediction error types across manually reviewed notes.

| Label | Label Definition | Percentage (%) |
| --- | --- | --- |
| 0 | Not medication ground truth | 2.631 |
| 1 | Accurate medication type, but non-specific | 5.263 |
| 2 | Accurate medication treatment, but different class | 15.789 |
| 3 | Wrong medication treatment, but appropriate in sentence context about disease | 39.473 |
| 4 | Wrong medication treatment, and not appropriate in sentence context about disease | 34.210 |
| 5 | Same medication but different name | 2.631 |

## DISCUSSION

Models trained on Medical ICU data outperformed those trained on Cardiothoracic or Surgical ICU data. This performance gap likely stems from a closer alignment between the Medical ICU training data and the test distribution, allowing the model to better learn relevant clinical patterns, terminology, and treatment norms. In contrast, the Cardiothoracic and Surgical ICUs represent narrower and more specialized patient populations, which may limit the generalizability of their models when applied to the more diverse cases typical of the Medical ICU.

Our study demonstrates the feasibility of developing ICU-specific digital twins using LLMs enhanced with LoRA adapters, focusing on medication prediction as a proof-of-concept task. Our findings underscore the superior performance of specialty-specific digital twins, such as the Medical ICU model, compared to both multi-disciplinary and other specialty models when predicting medications. This highlights the utility of tailored digital twins in augmenting decision-support systems and enhancing medication management in critical care.

Our findings suggest that while zero-shot approaches (akin to using general medical knowledge) have some utility, the supervised fine-tuning that incorporates specialty-specific practice patterns significantly enhances performance. This mirrors the clinical reality that ICU specialists develop expertise not just through textbook knowledge but through years of practice within their specific critical care environment.

The superior performance of specialty-specific digital twins, particularly the Medical ICU model, compared to both multi-disciplinary and other specialty models when predicting medications, highlights the importance of context-specific training. This performance differential is clinically significant, as medication management in the ICU requires precision tailored to the unique patient populations and practice patterns of each specialty. For example, a Medical ICU physician managing a patient with septic shock requires different medication considerations than a Cardiothoracic ICU physician managing a post-operative cardiac patient, despite both working within critical care environments.

The integration of digital twins into ICU workflow offers several advantages for specialists. First, by leveraging both the internal medical knowledge of LLMs and the real-world practice data embedded in EHRs, these systems can help alleviate the significant cognitive burden faced by ICU physicians who must process massive amounts of dynamic patient data while making time-sensitive decisions. Second, digital twins can serve as cognitive extensions that adapt to the specific knowledge base and practice patterns of different ICU specialties, potentially reducing medication errors and improving standardization of care.

The implications of digital twins extend beyond medication prediction. These systems can support dynamic, real-time simulations for monitoring and treatment planning, optimizing care in areas such as glycemic control, cardiovascular support, and mechanical ventilation.^28,29^ Digital twins additionally support the automation of care workflows, improving operational efficiency while addressing human factors to ensure effective usability in the dynamic ICU setting. Successful implementation relies on achieving robust interoperability among medical devices and ongoing research aimed at refining model accuracy and adaptability to the complexities of critical care environments^28^. While it is conceivable that a digital twin can be created via crafting a prompt that explicitly states the purpose and the specifications of the digital twin, we find that supervision is critical to the creation of digital twins. The reasoning process of physicians incorporates (1) knowledge representation in medicine, typical of medical board exam questions and textbooks; (2) practice experience represented by years of clinical training at the bedside, and (3) familiarity with best practices within an individual ICU and health system. Incorporating the EHR via adapters brings in many years of practice experience recorded in the documentation of EHR care notes within an ICU specialty and adds unique knowledge to the LLM. The local practice behaviors are also represented in the EHR as health systems have their own unique formulary of medicines available and guidelines such as antibiograms to follow. Our use of LoRA across multiple medical specialties allows the flexible interchange of LLMs to represent multiple digital twins and allow easy updating of the model as more EHR data arrives.

Finally, our work highlights the limitations of current metrics for evaluating generative models. Balancing the need to account for semantic variability in language with the risk of being overly permissive necessitates using multiple metrics, as no single metric provides a comprehensive assessment of generative performance. Exact-match metrics like accuracy and ROUGE-L are likely to underestimate performance (as indicated by our error analysis), while soft-match metrics like BERTScore may overestimate it, particularly for long sequences. At the expense of a more stringent, straightforward evaluation such as single medication prediction, our task was more clinically relevant to use document-level input from the EHR and generate all relevant medications. Our iterative approach refined the evaluation framework, settling on clinically relevant tasks that better reflect real-world applications. This approach revealed sensitivity in model performance, such as the impact of repeated medication mentions and long sequences on prediction accuracy, emphasizing the need for robust and context-sensitive evaluation methods.

One limitation of this study is that the models were trained exclusively on the MIMIC-III dataset, which represents a single-center ICU population. As such, the findings may not fully generalize to other hospital systems with differing patient demographics, clinical practices, or documentation styles. Additionally, because evaluation was performed only on Medical ICU notes, the performance of the Cardiothoracic and Surgical ICU models within their own respective domains remains untested. While our results support the effectiveness of specialty adaptation for the Medical ICU, further validation is needed to confirm the utility of this approach across other ICU specialties. Future work will evaluate each specialty-specific model on domain-aligned datasets and explore training and evaluating models on multi-institutional datasets to better assess cross-site generalizability and robustness.

## CONCLUSION

In this study, we presented an ICU specialty-adapted approach to digital twin modeling using LLMs, focusing on the task of medication prediction. Our results show that fine-tuning general-purpose LLMs with Low-Rank Adapters (LoRA) significantly improves their performance on specialty-specific clinical text, capturing nuanced differences in treatment patterns across ICUs. This demonstrates that even lightweight adaptation techniques can enable LLMs to serve as effective digital twins, mirroring the decision-making behaviors of specific ICU environments.

The ICU specialty adaptation plays a critical role in enhancing the relevance and accuracy of predictions, highlighting the importance of contextual grounding in clinical decision support. Rather than a one-size-fits-all model, our results support the development of modular digital twins, each tuned to the unique characteristics of a clinical domain.

In future work, we will investigate real-time model adaptation and expand the scope of prediction tasks beyond medications to include procedures, labs, and event forecasting. We also aim to explore digital twin alignment with individual healthcare providers to better personalize decision support at both the team and clinician level. These directions represent promising next steps toward building truly interactive, adaptive decision-support systems for critical care medicine.

## Data Availability

All data produced in the present study are available upon reasonable request to the authors

## AUTHOR CONTRIBUTIONS

The study was conceptualized by all authors. **Dmitriy Dligach** and **Majid Afshar** were responsible for designing the methodology. **Behnaz Eslami** implemented and further developed the methodology. The analysis of the results and the initial drafting of the manuscript were carried out by **Dmitriy Dligach, Majid Afshar**, and **Behnaz Eslami. Mathew Churpek** secured funding. All authors contributed to the review and revision of the manuscript, ensuring the accuracy and integrity of the work.

## SUPPLEMENTARY MATERIAL

Supplementary material is available at Journal of the American Medical Informatics Association online.

## FUNDING

This study received funding from the National Heart, Lung, and Blood Institute, United States, under Grant ID NIH 1R01HL157262, and the U.S. National Library of Medicine, United States, under Grant ID NIH R01 LM012973.

## CONFLICT OF INTEREST

The authors declare that they have no known competing financial interests or personal relationships that could have appeared to influence the work reported in this paper.

## DATA AVAILABILITY

It is available upon request.

## Appendix

**Table S1.** Error analysis of SparkNLP-based NER for clinical medication extraction.

| Metric | DRUG | DOSAGE | STRENGTH | ROUTE | FREQUENCY | DURATION | FORM |
| --- | --- | --- | --- | --- | --- | --- | --- |
| **Precision** | 0.946 | 0.79 | 1 | 0.958 | 0.944 | 0.944 | 0.833 |
| **Recall** | 0.939 | 1 | 0.982 | 0.983 | 0.944 | 1 | 1 |
| **F1_Score** | 0.942 | 0.883 | 0.991 | 0.971 | 0.944 | 0.971 | 0.909 |

**Table S2.**
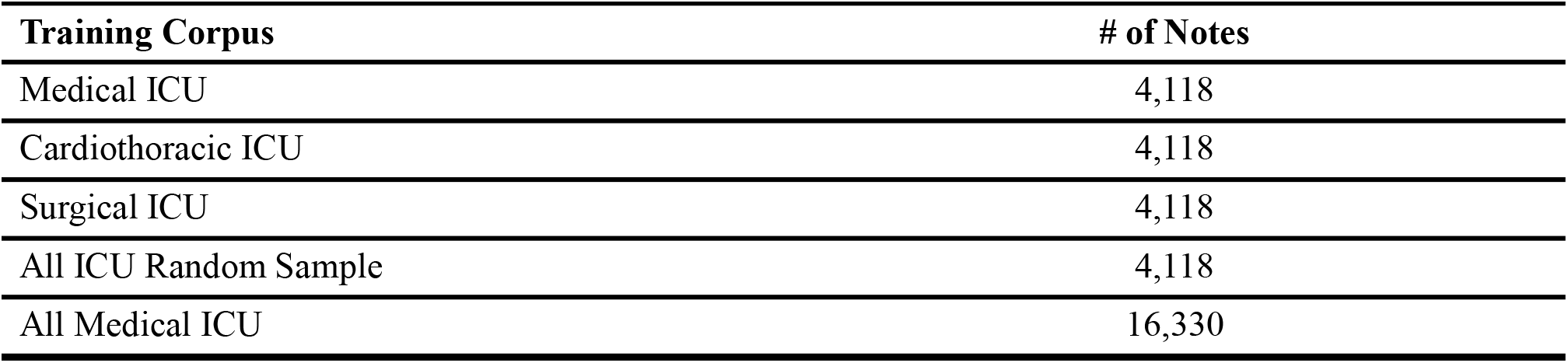
Distribution of training sets across various ICU types.

**Table S3.** Number of notes in test and validation sets for medical ICU only.

| Set Type (Only Medical ICU) | # of Notes |
| --- | --- |
| Validation Set | 1,500 |
| Test Set | 1,000 |

**Table S4.** Demographic and Clinical Characteristics of the Patient Cohort. The table presents the distribution of patients by sex, race/ethnicity, and primary diagnosis, reported as counts and corresponding percentages.

| Patient Characteristics | Size (Percentage) |
| --- | --- |
| **Sex, n (%)** |  |
| Female | 9,588 (43.425) |
| Male | 12,491 (56.574) |
| **Race/Ethnicity, n (%)** |  |
| White | 24,966 (81.079) |
| Black/African American | 3,739 (12.142) |
| Hispanic/Latino | 915 (2.971) |
| Other | 693 (2.250) |
| Asian | 479 (1.555) |
| **Diagnosis, n(%)** |  |
| Essential Hypertension | 13887 (27.4) |
| Congestive Heart Failure Unspecified | 10560 (20.9) |
| Atrial Fibrillation | 9535 (18.8) |
| Coronary Artery Disease (CAD) | 8626 (17.1) |
| Acute Kidney Failure | 7901 (15.6) |

**Table S5.**
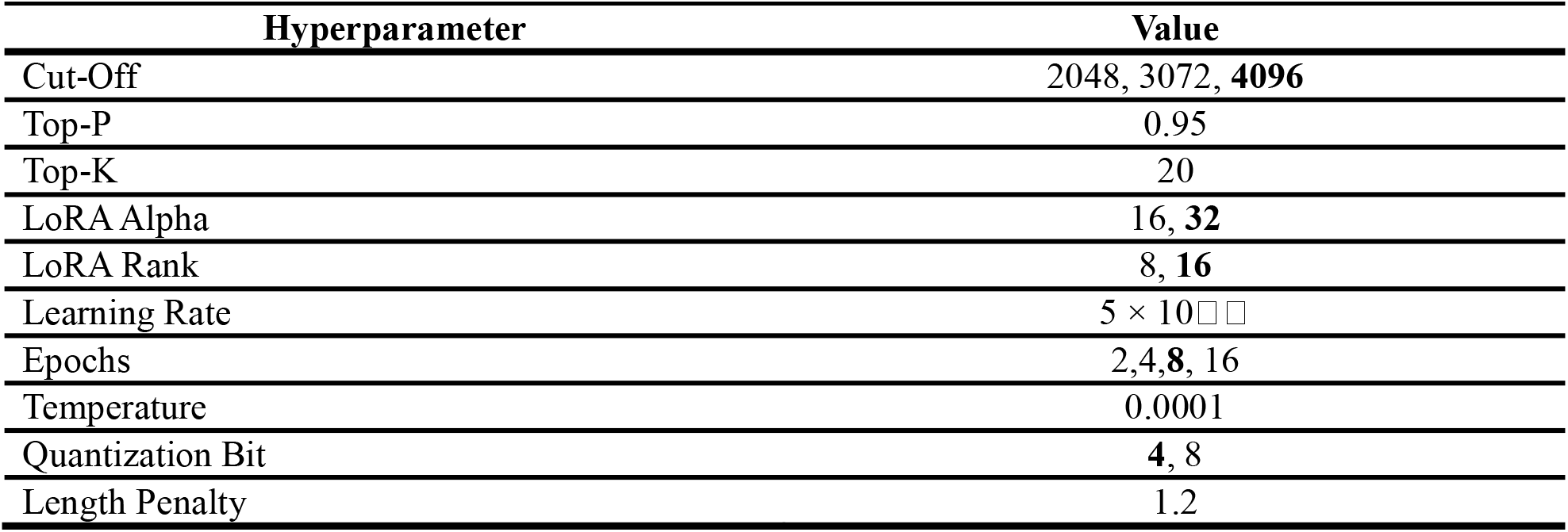
Evaluated hyperparameter values with the highlighted optimal values. The optimal values, determined through extensive evaluation, are bolded to indicate the best configuration for model training.

## Notes

### Competing Interest Statement

The authors have declared no competing interest.

### Author Declarations

Loyola University Chicago Institutional Review Board (IRB) reviewed and determined that the research project titled 'Learning Patient Representations from Electronic Health Records' is exempt from IRB oversight requirements, according to 45 CFR 46.101, as of February 15, 2024.

### Summary of Updates

This version of the manuscript has been thoroughly revised following both major and minor rounds of review. Key updates include clarification of the scope and framing of the study. Specifically, we now explicitly state that our model does not incorporate real-time, patient-specific EHR data, and we describe our approach as an ICU specialty-level language model adaptation rather than a fully realized digital twin. The Methods section has been expanded to include detailed descriptions of data sources, distribution of ICU types, data splitting strategies, and patient demographic characteristics. We clarified that the dataset split was performed at the patient level with negligible overlap. A new visual representation of the data flow has also been added to improve clarity. We provided additional implementation details, including key hyperparameters used during model training and evaluation. The validation results for the clinical NER model have been included, along with an error analysis of model predictions to highlight common failure patterns and their clinical context. The Discussion and Conclusion sections were revised to better highlight the value of fine-tuning with Low-Rank Adapters (LoRA) in improving the model's predictive performance. We emphasized how ICU specialty adaptation enhances clinical relevance and improves structured prediction tasks in domain-specific settings. We also addressed limitations, including generalizability concerns due to training on a single-center dataset, and described plans for evaluating the approach on multi-center data in future work. Structural and language improvements were made throughout the manuscript to improve readability and organization. We also confirmed that the implementation code and additional resources will be made publicly available upon acceptance. This updated version reflects all requested changes and presents the finalized manuscript in a more transparent, rigorous, and accessible format.

## REFERENCES

1. Grieves, M. & Vickers, J. Digital twin: Mitigating unpredictable, undesirable emergent behavior in complex systems. Transdisciplinary perspectives on complex systems: New findings and approaches 85–113 (2017).

2. Pellegrino, G., Gervasi, M., Angelelli, M. & Corallo, A. A Conceptual Framework for Digital Twin in Healthcare: Evidence from a Systematic Meta-Review. Information Systems Frontiers 1–26 (2024).

3. Vallée, A. Digital twin for healthcare systems. Front Digit Health 5, 1253050 (2023).

4. Schwartz, S. M., Wildenhaus, K., Bucher, A. & Byrd, B. Digital twins and the emerging science of self: implications for digital health experience design and “small” data. Front Comput Sci 2, 31 (2020).

5. Johnson, A. E. W. et al. MIMIC-III, a freely accessible critical care database. Sci Data 3, 1–9 (2016).

6. Batty, M. Digital twins. Environment and Planning B: Urban Analytics and City Science vol. 45 817–820 Preprint at (2018).

7. Jiang, Y., Yin, S., Li, K., Luo, H. & Kaynak, O. Industrial applications of digital twins. Philosophical Transactions of the Royal Society A 379, 20200360 (2021).

8. of Sciences Engineering, Medicine & others. Foundational Research Gaps and Future Directions for Digital Twins. (2023).

9. Meta, A. I. Introducing meta llama 3: The most capable openly available llm to date, 2024. URL https://ai.meta.com/blog/meta-llama-3/. Accessed on April 26, (2024).

10. Hu, E. J. et al. Lora: Low-rank adaptation of large language models. arXiv preprint arXiv:2106.09685 (2021).

11. Venkatesh, K. P., Raza, M. M. & Kvedar, J. C. Health digital twins as tools for precision medicine: Considerations for computation, implementation, and regulation. NPJ Digit Med 5, 150 (2022).

12. Achiam, J. et al. Gpt-4 technical report. arXiv preprint arXiv:2303.08774 (2023).

13. Touvron, H. et al. Llama: Open and efficient foundation language models. arXiv preprint arXiv:2302.13971 (2023).

14. Singhal, K. et al. Large language models encode clinical knowledge. Nature 620, 172–180 (2023).

15. Nazi, Z. A. & Peng, W. Large language models in healthcare and medical domain: A review. in Informatics vol. 11 57 (2024).

16. Liu, Q. et al. Large language model distilling medication recommendation model. arXiv preprint arXiv:2402.02803 (2024).

17. Dou, Y. et al. ShennongGPT: A Tuning Chinese LLM for Medication Guidance. in 2023 IEEE International Conference on Medical Artificial Intelligence (MedAI) 67–72 (2023).

18. Ahmed, A., Zeng, X., Xi, R., Hou, M. & Shah, S. A. MED-Prompt: A novel prompt engineering framework for medicine prediction on free-text clinical notes. Journal of King Saud University-Computer and Information Sciences 36, 101933 (2024).

19. Aden, I., Child, C. H. T. & Reyes-Aldasoro, C. C. International Classification of Diseases Prediction from MIMIIC-III Clinical Text Using Pre-Trained ClinicalBERT and NLP Deep Learning Models Achieving State of the Art. Big Data and Cognitive Computing 8, 47 (2024).

20. Kocaman, V. & Talby, D. Improving clinical document understanding on COVID-19 research with spark NLP. arXiv preprint arXiv:2012.04005 (2020).

21. Dubey, A. et al. The llama 3 herd of models. arXiv preprint arXiv:2407.21783 (2024).

22. Liu, F., Shareghi, E., Meng, Z., Basaldella, M. & Collier, N. Self-alignment pretraining for biomedical entity representations. arXiv preprint arXiv:2010.11784 (2020).

23. Bodenreider, O. The unified medical language system (UMLS): integrating biomedical terminology. Nucleic Acids Res 32, D267–D270 (2004).

24. Croxford, E. et al. Development of a Human Evaluation Framework and Correlation with Automated Metrics for Natural Language Generation of Medical Diagnoses. medRxiv (2024).

25. Guo, X. & Vosoughi, S. Length does matter: Summary length can bias summarization metrics. in Proceedings of the 2023 Conference on Empirical Methods in Natural Language Processing 15869–15879 (2023).

26. Zhang, T., Kishore, V., Wu, F., Weinberger, K. Q. & Artzi, Y. Bertscore: Evaluating text generation with bert. arXiv preprint arXiv:1904.09675 (2019).

27. Lin, C.-Y. Rouge: A package for automatic evaluation of summaries. in Text summarization branches out 74–81 (2004).

28. Geoffrey Chase, J. et al. Digital twins and automation of care in the intensive care unit. Cyber– Physical–Human Systems: Fundamentals and Applications 457–489 (2023).

29. Thangaraj, P. M., Benson, S. H., Oikonomou, E. K., Asselbergs, F. W. & Khera, R. Cardiovascular care with digital twin technology in the era of generative artificial intelligence. Eur Heart J ehae619 (2024).

